# The SARS-CoV2 pandemic explained via asymptomatic infection and susceptibility heterogeneity. Buenos Aires first wave

**DOI:** 10.1101/2021.04.26.21255801

**Authors:** Manuela Bullo, Santiago Poy-Piñeiro, Pedro Cosatto-Ammann, Hernan Seoane, Emilio Picasso

**Author notes:** Manuela Bullo is the corresponding author. Manuela Bullo. Santiago Poy-Piñeiro, Pedro Cosatto Ammann, Hernan Seoane, Emilio Picasso.

## Abstract

**Background:** The rapid global spread of SARS-COV-2 forced governments to implement drastic interventions. The existence of a large but undetermined number of mild or non-symptomatic but infectious cases seems to be involved in the rapid spread, creating a high level of uncertainty due to the difficulty to measure them, and difficulty for epidemiologic modelling.

**Methods:** We developed a compartmental model with deterministic equations, that accounts for clinical status, mobility, r heterogenous susceptibility and non-pharmaceutical interventions. The model was calibrated using data from different regions and we used it to predict the dynamic in Buenos Aires Metropolitan Area (AMBA).

**Results:** The model adjusted well to different geographical regions. In AMBA the model predicted 21400 deaths at 300 days, with 27% of the population in the region immunized after the first wave, partly due to the high incidence of asymptomatic cases. The mobility restriction is approximately linear, with any restriction bringing a positive effect. The other interventions have a combined effect of 27% reduction in infection rates.

**Conclusion:** Our research underlines the role of asymptomatic cases in the epidemics’ dynamic and introduces the concept of susceptibility heterogeneity as a potential explanation for otherwise unexplained outbreak dynamics. The model also shows the big role of non-pharmaceutical interventions both in slowing down the epidemic dynamics and in reducing the eventual number of deaths. The model results are closely compatible with observed data.

## Background

At the end of 2019, a new coronavirus was identified in Wuhan, China, with rapid global spread. Different mitigation and contention strategies have been used globally, despite the lack of clear evidence of their impact and relevance. Mathematical models provide a framework to understand the epidemics dynamics in this changing setting and to plan and evaluate different control measures.

A wide range of mathematical models have been proposed. Most models use the SIR – SEIR framework with different variants, formulated in terms of ordinary differential equations (1) (2) Based on Ross’ theory of dependent happening (3) (4), they assume that incidence of infectious diseases at time *t* depends on prevalence of the disease, contact rate and transmission probability per contact. In other words, epidemiologists model transmission as a dynamic process, with variables changing in response to themselves. There are two phases in this type of process: adjustment and prediction. In the first phase, using data from the past, the model “learns” some parameters, and then, in the second phase, it uses these learned parameters to predict evolution of the epidemics in a different setting.

During COVID-19 pandemic, mathematical models have flourished trying to answer a demand from decision makers and society in general. Early in March 2020, Shaman et al estimated with their model the role of substantial undocumented infection in the transmission of the disease (5), proposing that 86% of infections were undocumented, and accounted for 79% of transmissions in Wuhan, China. The Imperial College of London published an influential report in March 2020, underscoring the importance of combined non-pharmaceutical interventions to contain the spread of the virus, for as long as 18 months (6). While the previously mentioned models estimated an R_0_ between 2 and 3, Sanche et al (7) estimated even higher R_0_ than previous models, sustaining the need of combined strategies to contain the virus. Ivorra’s group at Universidad Complutense de Madrid propose a SEIR model with 9 compartments, including hospitalized patients that die separate from hospitalized patients that live (8) (9). Mordecai’s group at Stanford University developed a model that includes asymptomatic individuals (10).

Globally, governments implemented strict responses (lockdowns, social distancing, school closing and restrictions in social gatherings) during the disease outbreak (11). These responses had a relevant effect on the reduction of Covid-19 incidence (12) and made mathematical modelling more complex since models now have to account for several governmental measures and changes in social behavior (13). Also, the quality of the parameters estimated by the models depends widely on the data used to adjust the parameters. Due to scarcity and low quality of data that have been reviewed in real-time during the COVID pandemic, some parameters had to be re-estimated once and again, and some models have been widely questioned due to lack of precision and ever-changing predictions. On the other hand, these models have influenced health decisions worldwide, and have been critical in the evolving dynamic of this pandemic.

With the goal of developing a model that reflects the epidemics in Argentina with its epicenter in Buenos Aires and its suburban area, we developed a new model, innovative in that it accounts for heterogeneous susceptibility, mobility index and clinical status. We calibrated it to outbreaks in other relevant areas (Spain and the New York State) and then used it to assess different scenarios of the epidemics in Buenos Aires Metropolitan Area (AMBA), particularly the governmental responses taken to mitigate the spread of the disease.

## Methods

### Data sources

The model is focused on the metropolitan area of Buenos Aires (AMBA), which comprises the Autonomous City of Buenos Aires and 24 adjacent districts in the Province of Buenos Aires, has an area of 3.830 km^2^ and, according to the last Census, 12.8 million inhabitants (almost a third of the national population). The proposed model required different data sources: reported cases and deaths due to SARS-CoV-2 disease, estimations of the proportion of underreported symptomatic cases, and, finally, data of government responses to the COVID-19 outbreak and changes in people mobility.

Since the AMBA is a socioeconomic agglomeration but its boundaries do not correspond to political or administrative borders, a highly disaggregated dataset was required in order to assign confirmed cases and deaths to their specific residential area. Data for AMBA was prepared on the basis of an open dataset of the National Health Ministry (*Ministerio de Salud*), which is compiled online by the authorities through the National Service of Health Surveillance and data is entered manually at the point of care (14). It is an open public dataset that includes a disaggregated urban code, with basic demographic information, confirmed cases and deaths and it is updated daily at 5.45 PM (UTC-3).

The proportion of underreported cases were estimated for several countries by the Centre for Mathematical Modelling of Infectious Diseases (CMMID) of the London School of Hygiene & Tropical Medicine (15), assuming that not all the cases are being detected. According to Russell et al., the methodology has two main steps. Firstly, they correct the naïve case fatality ratio (nCFR) –that is, the ratio of reported deaths to reported cases to date– which is underestimated due to the delay to the proper outcome (recovery or death) and obtain a corrected CFR (cCFR). Secondly, by using previous research of China and South Korea on the case fatality ratio, they calculate for every country an underreporting level on a daily basis. The calculation performed is .014/cCFR, where .014 is the case fatality ratio as informed by several large studies and cCFR is the corrected CFR aforementioned. Underreporting is assumed to stabilize after the period reported.

Furthermore, our model includes a contact function that required data on public responses to the pandemic and people mobility changes since the outbreak started. We used data on government responses from the Oxford COVID-19 Government Response Tracker(11), which includes different indicators of the responses adopted, measured in a scale of stringency. Specifically, our contact function includes information of school closing, cancelation of public events and the severity of the restrictions on gatherings.

Finally, the model requires data on mobility trends, which are provided publicly by Google through their Mobility Report dataset (16). This dataset shows how visits and lengths of stay at different places changed compared to a baseline, using data from users Location History of their Google Account. This baseline is the median value for the corresponding day of the week during a five-week period between January 3^rd^ and February 6^th^. We used changes in mobility patterns in places defined as public transport stations (such as underground, bus and train stations) and workplaces. With data for AMBA, we elaborated an index of mobility changes that considers the population size of each department of the AMBA as a weighting factor.

During the adjustment stage of the model, we used data from areas that faced an earlier outbreak: Spain, a country which is culturally tied to Argentina, and the State of New York, a large metropolitan area that experienced a rapid spread of the disease. Data of reported cases and deaths for Spain were taken from the World Health Organization (WHO) *Coronavirus Disease (COVID-19) Dashboard* dataset (17). The Health Ministry (*Ministerio de Sanidad*) (18) carried out a data reconciliation that implied removing a large number of confirmed cases and deaths from the total count. We have corrected negative numbers between May 26^th^ and June 5^th^by reassigning classification errors proportionally to total cases (or total deaths) in days prior to those dates. As for the State of New York, we used data from the Coronavirus Resource Center of the Johns Hopkins University (19).

### The Model

We have designed the deterministic compartment model shown in Figure 1. The S compartment holds the susceptible individuals that gradually become exposed to the virus and move to the E compartment. Exposed individuals become infected and may develop symptoms (IS) or not (IA). This process is controlled by the parameters: θ_S_ (fraction of symptomatic) and γ_E_ (inverse of the incubation period). Symptomatic cases may be severe (HS), moderate (HM) or mild (IL), following the fractions θ_HS_, θ_HM_, and (1- θ_HS_ - θ_HM_) respectively, and the rate γ_IS_. Both severe and moderate cases are confirmed and isolated. Severe cases are hospitalized and require ICU, that may or may not be available. Some of them die (D) at the rate γ_HSD_, and the other recover at a specific rate: γ_HSR_. The fraction of severe cases that die: ω_HS_ is introduced in the model as a decreasing function of time reflecting the learning curve as found by Cook (2020) (20) and subsequent improvements (WHO, 2020) (21). Thus, the fatality rate among symptomatic is ω = θ_HS_ω_HS_. Moderate cases are either hospitalized (not in ICU) or isolated at home, and they are assumed to recover at a rate γ_HM_. Mild symptom cases remain undetected and recover at the rate γ_IL_. On top of that, the model allows for completely asymptomatic cases that contribute to the infection process, and eventually recover (stop transmitting the disease) at a rate γ_IA_.

**Figure 1:**
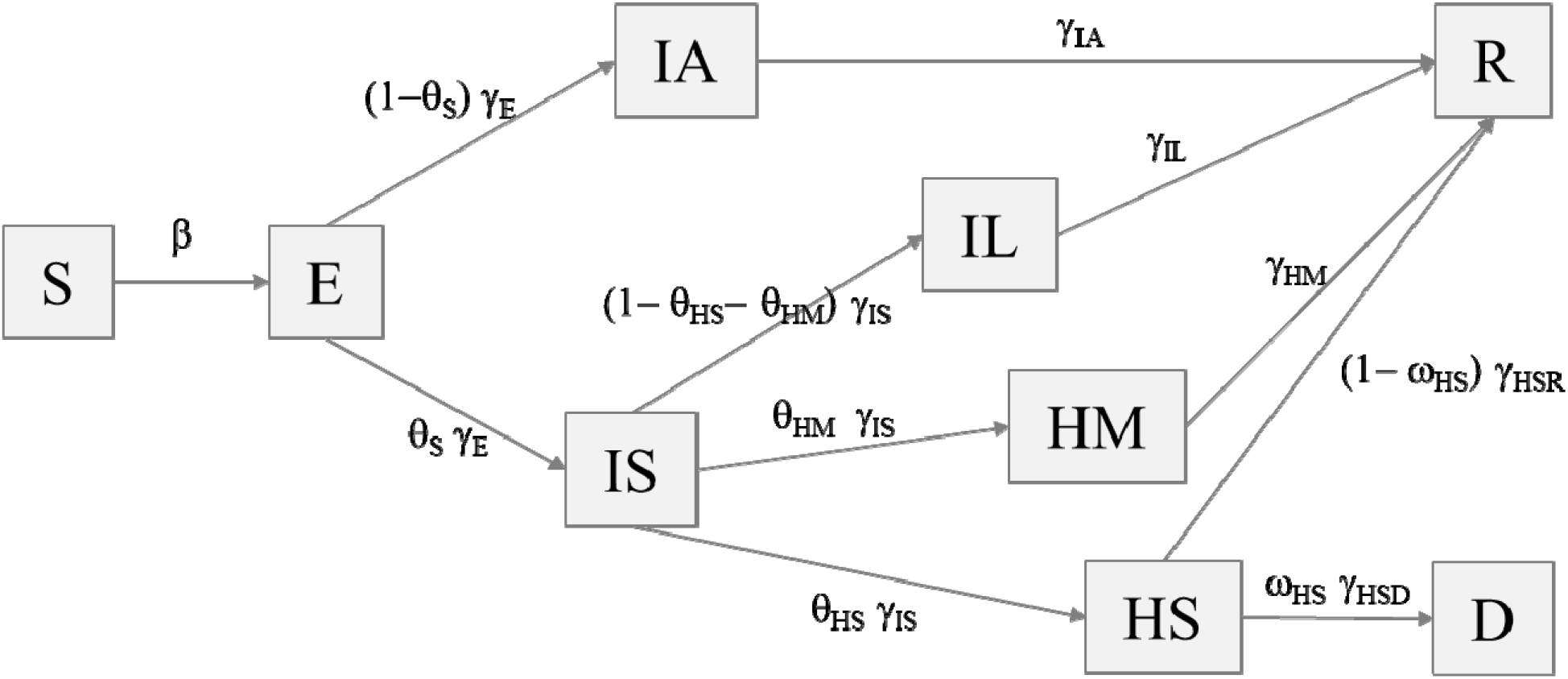
Proposed model. Source: Authors’ own elaboration.

The infection process is produced by function (1), introducing the contagion parameters (beta) from all infectious compartments:

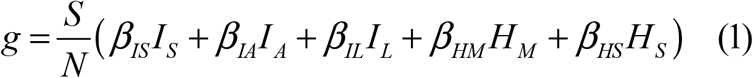

Where *N* stands for the number of habitants in the population, assumed constant throughout the epidemic. Natural birth-death process is considered negligible given the fast development of the SARS-CoV2 epidemic. Import-export process is also discarded given the travel restrictions established by the governments, except for the initial situation. The early confirmed cases are sown backwards as exposed individuals until the number of imported cases reported by the local government is completed, for the model to shift them downstream to the subsequent stages.

The system of differential equations of the model follows from the model diagram:

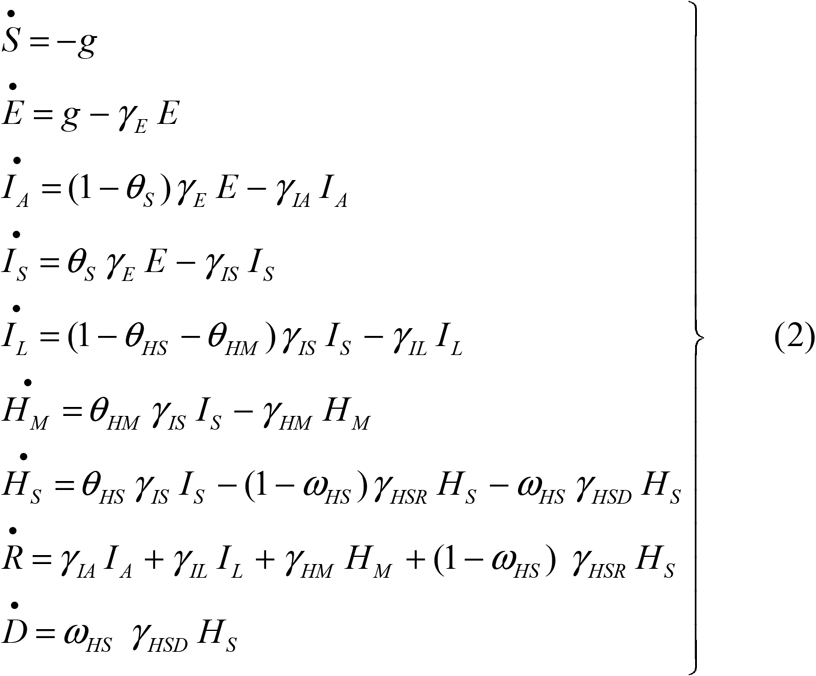

The number of severe hospitalized cases is limited by the capacity of the health system (number of ICU beds). The excess cases move to the D compartment as they occur, assuming that all severe cases requiring but not getting ICU will die immediately.

The gamma parameters, mostly associated to natural phenomena, are obtained from previous research or, when it is possible, from data for Argentina, as shown in table 1.

**Table 1.**
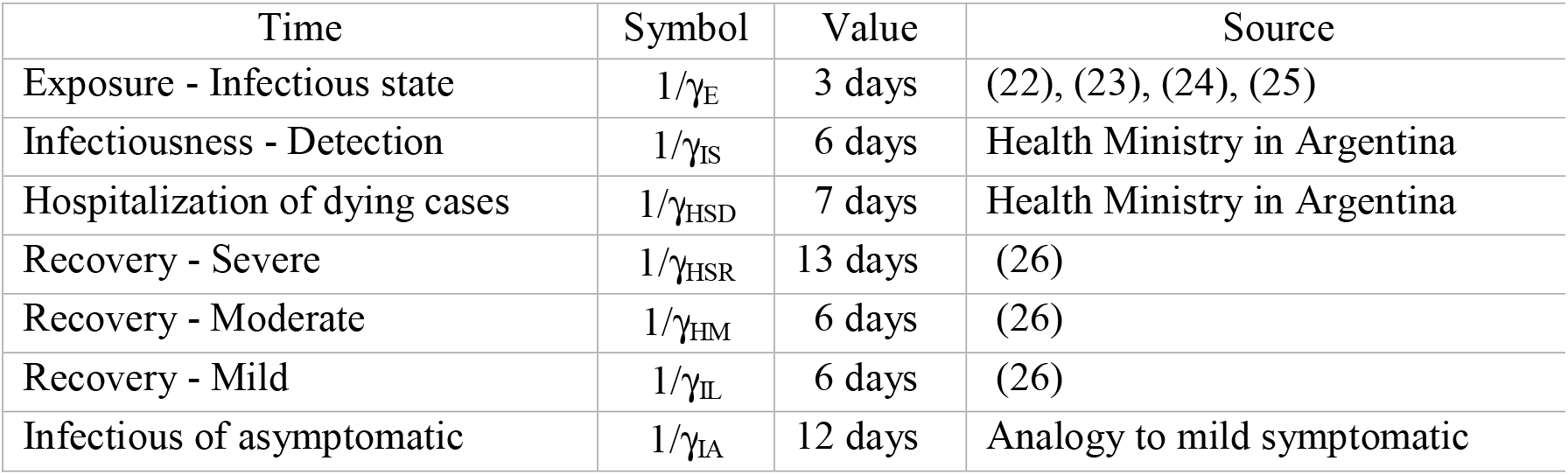
Gamma Parameters.

The beta and theta parameters are estimated to fit the data of daily deaths and cases in the target population. The symptomatic cases are calculated out of confirmed cases by means of the underreporting methodology described above. The differential equations (2) are discretized for recurrent solution. The error functional follows the minimum squares criterion, and the optimization is performed via simulated annealing to take into account its complexity and potential multiple local optima.

The intervention established by the governments is modeled within the beta parameters. We propose the following function (3) to liaise the intensity of the intervention and the number of contacts:

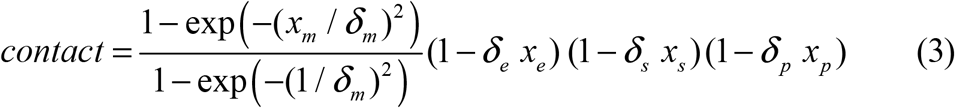

The first factor models the influence of mobility restrictions on the infection parameters. We operationalize the intensity of this intervention by means of the mobility index produced by Google (*x*_*m*_), specifically the average of the transit and workplaces indexes as previously explained(16). In this way we bring into the model not only the government decisions, but also the shortcoming behavior of uncompliant individuals, and the overreaction of individuals self-restricting beyond the authority regulations. This function has the ability to adapt to different forms, depending on δ_m_, as shown in Figure 3. On one side the combinatorial (quadratic) case where every person moving meets each other is represented with large δ_m_. Any slight intervention would bring a positive effect in this case. On the other side, saturated forms where only very intense interventions would have a noticeable effect are obtained with low values of δ_m_. The parameter δ_m_ is estimated in the model calibration.

**Figure 3:**
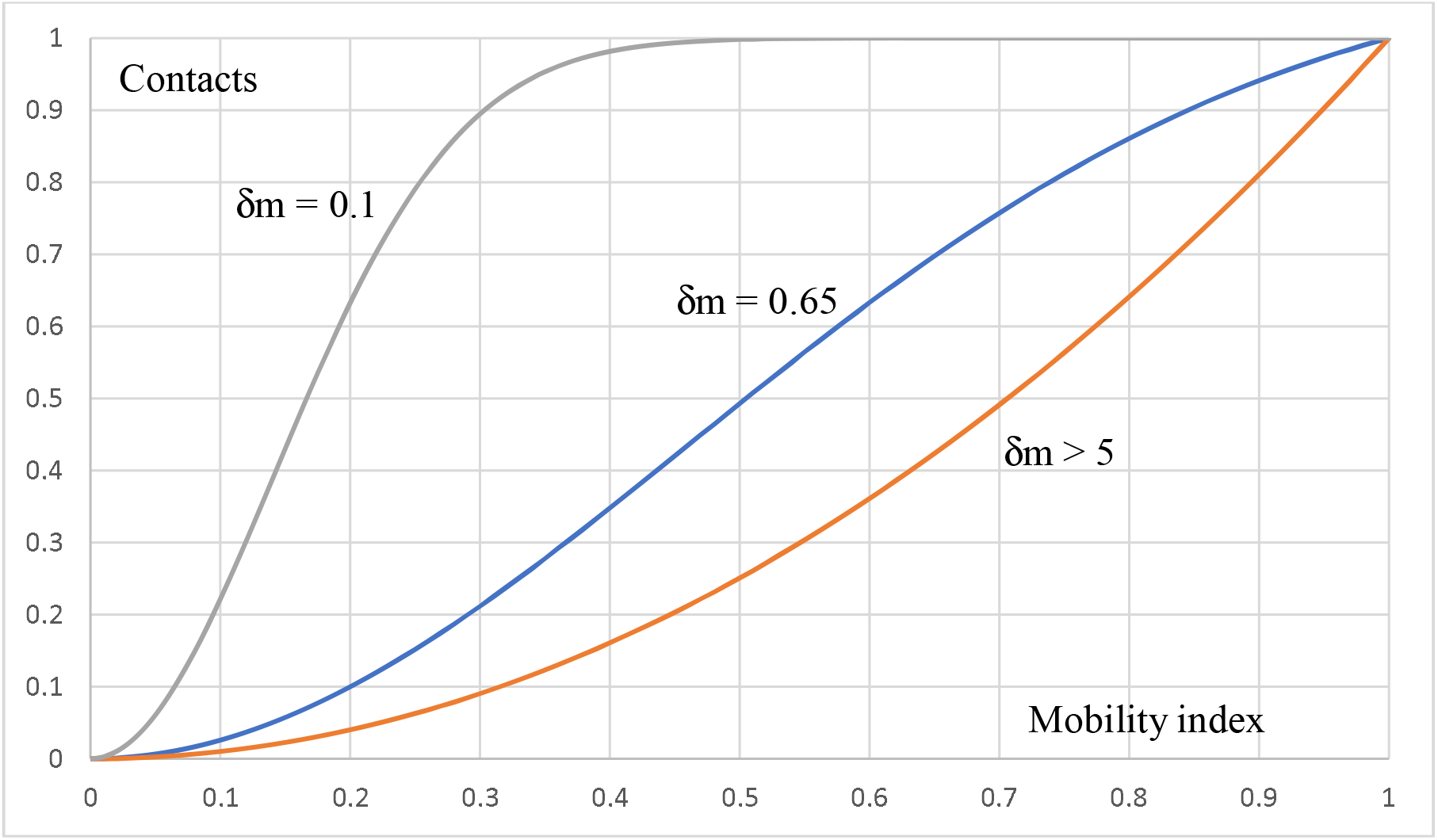
Contact mobility function. Source: Authors’ own elaboration.

While the effect of gatherings is indirectly considered in the mobility index, as gathering implies movement, the act of gathering itself would likely bring additional infection. Then, we model this increased effect by means of two binary variables entering as second and third factors: the prohibition of massive events and gatherings (*x*_*e*_), and the suspension of school and university activity (*x*_*s*_). Another element contributing to slow down the proliferation of the disease is the precautionary behavior of the population, induced by the communication from the government and the media. We operationalize this in the fourth factor by means of a binary variable indicating the obligation to use masks (*x*_*p*_), that stands out as one of the most spectacular government directives strongly inducing behavior change. The delta parameters measure the effect of the interventions on the infection beta parameters, and they are estimated together with them in the model calibration phase. The beta parameters of non-isolated individuals (IS, IA, IL) in expression (1) are affected by the contact function (3), hence the estimated beta parameters represent the maximum values prevailing in the absence of intervention.

The issue of susceptibility is currently under discussion. Differences in lethality have been identified due to a series of risk factors like sex, age, and a series of comorbidities. Beyond that, susceptibility to infection can be heterogeneous due to biological factors, like cross-immunity with other coronaviruses (27), T-cell based immunity in seronegative individuals (28), different viral load (29), and to geographical and behavioral factors, among others. The effect of heterogeneous susceptibility is introduced into the model by means of function (4) that indexes the probability of infection to the remaining susceptible population.

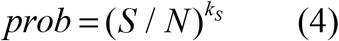

The assumption is that highly susceptible individuals are infected at the beginning of the epidemic, causing a reduction in the susceptibility of the remaining susceptible population. Function (4) enters the infection function (1) as a factor, hence the estimated beta parameters represent the initial values with maximum prevailing susceptibility.

## Results

The model is applied to different populations with different timing and intervention policies, for calibration stability. In all cases the model successfully reproduces the real evolution of symptomatic cases and deaths. The model is first applied to New York, Spain, and Sweden. The epidemic in these regions has completed the first outbreak by the time of writing, and they followed different intervention approaches. The learning from these cases is applied to the case of Buenos Aires, where the epidemic started by March 3^rd^ with the first confirmed case, it was contained by means of a strict and long intervention, and thus developed slowly and peaked by end of August. The first three cases are discussed in the appendix.

The results of the model are shown in figure 4. The model reproduces the real deaths and symptomatic cases evolution quite well. The peak of around 7900 daily symptomatic cases (adjusted for underreporting) is predicted by the end of August, and the peak of 185 daily deaths 10 days later. The total number of deaths during 300 days is predicted at 21400, assuming continued intervention.

**Figure 4:**
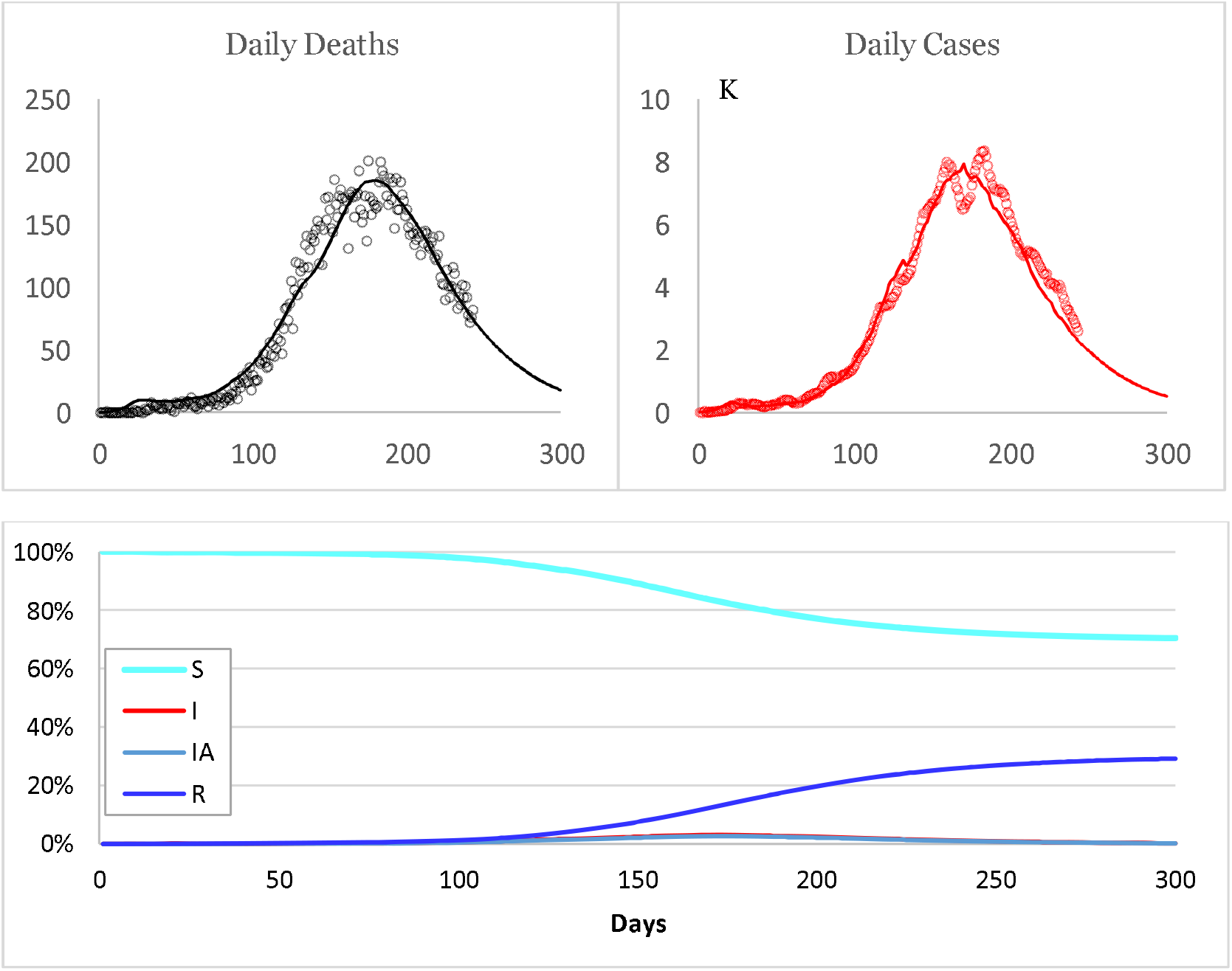
Model prediction for Buenos Aires Metropolitan Area. Source: Authors’ own elaboration.

The estimated parameters of the model are shown in table 2. The model predicts a numerous asymptomatic population (1-θ_S_ = 81%) having non negligible infectious activity (about 1/3 of symptomatic). This is compatible with empirical findings. The severe cases are estimated at 5.5% of symptomatic cases. The susceptibility distribution is convex (k_S_ = 2.75), reflecting a large core of low susceptibility population due to either biological or behavioral reasons. The mobility restriction is approximately linear, meaning that any restriction brings a positive effect. The other interventions have a combined effect of 27% reduction in infection rates. As shown in Figure 4, 29% of the population is estimated to be immunized after the epidemic wave, mainly due to the large asymptomatic base.

**Table 2.** Parameters of the model.

| $\beta_{IS}$ | $\beta_{IA}$ | $\beta_{IL}$ | $\beta_H$ | $k_S$ | $\theta_S$ | $\theta_{HS}$ | $\delta_m$ | $\delta_e$ | $\delta_s$ | $\delta_p$ |
| --- | --- | --- | --- | --- | --- | --- | --- | --- | --- | --- |
| 0.86 | 0.29 | 0.58 | 0.09 | 2.75 | 0.19 | 0.055 | 0.58 | 0.09 | 0.06 | 0.15 |

Source: Authors' own elaboration.

The model predicts 60000 deaths in the laissez faire scenario (no intervention throughout the whole period of observation), reflecting the value of the intervention.

## Discussion

COVID-19 has disrupted life worldwide. With the use of mathematical models, we have tried to understand the dynamic of the epidemics, and we propose a model that explains it, considering the non-pharmaceutical measures taken by governments and population in general. Our model predicts a significant fraction of asymptomatic cases developing immunity. This is compatible with empirical findings obtained through seroprevalence studies in the area (30; 31), considering T-cell based immunity. On the other end, over two thirds of the population would remain uninfected due to lower susceptibility. These results are sensitive to changes in mobility and flexibilization of non-pharmaceutical intervention.

Some strengths of our research can be pointed out. Our model explains the dynamics of the epidemic by means of two main forces, besides intervention. First, the large asymptomatic population, contributing to the rapid epidemic spread as much as to herd immunity. Second, heterogeneous susceptibility gradually slowing down the epidemic spread and eventually causing its extinction. This explanation also works for other regions analyzed in the appendix. We believe that this dual explanation allows the scientific community to raise new hypotheses about the dynamics of the outbreak contributing to better understanding. While we do not claim this one to be the definite explanation, our paper emphasizes the role of asymptomatic individuals in the spread of the outbreak, in accordance with increasing evidence in the field, and the role of susceptibility heterogeneity, highlighting areas for further research on this field. Different hypotheses could explain the susceptibility heterogeneity. On the biological side, some researchers have pointed out potential cross-immunity with other coronavirus, as well as other pre-existing health conditions. On the behavioral side, different levels of precautions could also explain heterogeneity. This would raise the possibility of new outbreaks as the precautions are relaxed.

As new epidemic outbreaks are appearing in several places, it would be interesting to understand the model capability to explain them. The phenomenon seems more intense in populations where the first outbreak did not widely spread, although it is not fully understood at the time that we write this article. The proposed model opens the possibility of new epidemic outbreaks as intervention of precautionary behavior relax. The model could be extended to better explain such phenomenon by introducing limited duration of immunity and pharmaceutical intervention.

Finally, our model is sensitive to mobility and non-pharmaceutical measures, allowing to predict different plausible scenarios according to the governmental measures and the population behavior. Nevertheless, the effect of the intervention predicted by the model is not massive. The number of lives saved in Buenos Aires is estimated at 38600.

There are naturally a series of simplifying assumptions in this model, like the asymptomatic or mild symptomatic cases detected and isolated on occasion of the investigation of the close connections of a confirmed case or a patient hospitalized for other reasons; the moderate or severe symptomatic cases that remain undetected; or the moderate symptomatic cases that die. However, we think that these cases are out of the mainstream of the epidemic, and they pose no serious limitation to the model. On the other hand, the proposed model is highly flexible, allowing for both completely asymptomatic and mild symptomatic cases, that are thought to play a significant role in the SARS-CoV2 epidemic.

Among the limitations of our research, we have to mention the data quality. Changes in real-time data due to corrections, poor data-quality and slow reporting may affect therefore the assumptions of our model. Under-reporting due to slow data processing, restrictive testing policies and lack of testing availability impacted on the cumulative number of cases acknowledged by official data sources. While we use Russell’s method to correct for this, we are introducing potential limitations of Russell’s method into our model. Also regarding data quality, imported cases also introduce uncertainty in the model, and data is not as granular as it is required to account for that. Another limitation is that in our model, 81% of the infections are asymptomatic. While as mentioned previously, some local data shows that for every PCR-diagnosed case there were 9 IgG SARS CoV-2 positive individuals that had not been diagnosed during the outbreak in a very poor neighborhood in the City of Buenos Aires (30) reaching 50-60% seroprevalence, studies conducted in Europe after local outbreaks show 15 to 20% seroprevalence (31). Further research is required to understand if this phenomenon is due to rapid decline in serum antibodies after asymptomatic infections or if asymptomatic individuals acquired non-antibody immunity, and differences in outbreaks and type of households and family composition may also play a role in this respect. Finally, we are not considering the role of the heterogeneous distribution of risk groups such as aging population in the different settings or the impact of extended households versus nursing homes. Further research should be conducted in these areas to understand their role in the outbreak dynamics.

In conclusion, our model highlights the importance of asymptomatic infection in the spread of this outbreak, is sensitive to changes in non-pharmaceutical measures and assumes that new outbreaks occur where past outbreaks were not strong enough to reach most population. The model assumptions are relaxed by including a susceptibility index that explains high speed spread at the beginning of the outbreak and reduction of cases while highly susceptible cases have recovered. According to our model, the interventions were useful to slow down the epidemic dynamics and to reduce the total number of deaths due to this disease. The model results are closely compatible with observed data, but due to the dubious quality of data that characterize this pandemic, it is difficult to say if it provides the true explanation. Further research, both empirical and modelling, is needed to help us understand more deeply this disease, while vaccines and pharmaceutical measures are being developed.

## Data Availability

All data used for this paper is available upon request

## Acknowledgements

We would like to acknowledge the support of Dr Miguel Angel Schiavone, Chancellor of the Catholic University of Argentina.

This research did not receive any specific grant from funding agencies in the public, commercial, or not-for-profit sectors.

## Data Availability

Raw data were generated at Catholic University of Argentina. Derived data supporting the findings of this study are available from the corresponding author MB on request.

